# A pilot dose-finding study of Terazosin in humans

**DOI:** 10.1101/2024.05.22.24307622

**Authors:** Jordan L. Schultz, Phillip E. Gander, Craig D. Workman, Laura L. Ponto, Stephen Cross, Christopher S. Nance, Christopher L. Groth, Eric B. Taylor, Sarah E. Ernst, Jia Xu, Ergun Y. Uc, Vincent A. Magnotta, Michael J. Welsh, Nandakumar S. Narayanan

## Abstract

**Background:** Parkinson’s disease (PD) is a prevalent neurodegenerative disorder where progressive neuron loss is driven by impaired brain bioenergetics, particularly mitochondrial dysfunction and disrupted cellular respiration. Terazosin (TZ), an α-1 adrenergic receptor antagonist with a known efficacy in treating benign prostatic hypertrophy and hypertension, has shown potential in addressing energy metabolism deficits associated with PD due to its action on phosphoglycerate kinase 1 (PGK1). This study aimed to investigate the safety, tolerability, bioenergetic target engagement, and optimal dose of TZ in neurologically healthy subjects.

**Methods:** Eighteen healthy men and women (60 – 85 years old) were stratified into two cohorts based on maximum TZ dosages (5 mg and 10 mg daily). Methods included plasma and cerebrospinal fluid TZ concentration measurements, whole blood ATP levels, ^31^Phosphorous magnetic resonance spectroscopy for brain ATP levels, ^18^F-FDG PET imaging for cerebral metabolic activity, and plasma metabolomics.

**Results:** Our results indicated that a 5 mg/day dose of TZ significantly increased whole blood ATP levels and reduced global cerebral ^18^F-FDG PET uptake without significant side effects or orthostatic hypotension. These effects were consistent across sexes. Higher doses did not result in additional benefits and showed a potential biphasic dose-response.

**Conclusions:** TZ at a dosage of 5 mg/day engages its metabolic targets effectively in both sexes without inducing significant adverse effects and provides a promising therapeutic avenue for mitigating energetic deficiencies. Further investigation via clinical trials to validate TZ’s efficacy and safety in neurodegenerative (i.e., PD) contexts is warranted.

## INTRODUCTION

Parkinson’s disease (PD) is the second most common neurodegenerative disease.^1, 2^ Current treatments can reduce some PD symptoms but no therapies prevent progressive neurodegeneration.^3^ However, recent research has demonstrated that impaired brain bioenergetics are a key factor in PD.^4^ Specifically, the mitochondrial toxin MPTP causes parkinsonism in humans and animal models of PD,^5–11^ and PD-causing mutations can disrupt cellular respiration.^12–19^ Furthermore, advancing age is the major risk factor for developing PD and is associated with disrupted energy metabolism,^20–26^ as is seen in the brains of PD patients.^27^ Therefore, interventions targeting energy metabolism have potential to slow or halt the progressive neuron loss seen in PD.^28^

Recent studies discovered that terazosin (TZ), an α-1 adrenergic receptor antagonist used to treat benign prostatic hypertrophy and hypertension, has an additional independent target in phosphoglycerate kinase 1 (PGK1).^29^ PGK1 is the first ATP-producing enzyme in glycolysis, and interestingly, human mutations in PGK1 are associated with Parkinsonism.^30–33^ The ability of TZ to enhance energy metabolism via activation of PGK1 suggested that it might improve impaired PD bioenergetics. In testing that hypothesis we found that TZ enhanced glycolysis and mitochondrial activity, increased ATP levels in the brain, and importantly, slowed or prevented neurodegeneration in diverse cellular and animal models of PD.^34^ Those discoveries were complemented by several large epidemiological studies that associated TZ use with a decreased risk of developing PD and a slower disease progression in people who already had PD.^35–39^ In addition, a small pilot study suggested that TZ engaged its target and increased ATP in people with PD.^40^

Although these findings are encouraging, there are still several key questions that need answers to better guide future clinical TZ utility and treatment strategies in people with PD. We aimed to fill these gaps in knowledge by studying the pharmacodynamic and pharmacokinetic properties of TZ in older healthy adults before moving to larger trials in patients with PD. First, does TZ cross the blood-brain barrier (BBB) in humans, enabling it to reach its intended target sites in the brain? Animal studies suggest that TZ penetrates the BBB^34^ but this remains largely unexplored in humans. Second, given that TZ is predominantly prescribed to treat benign prostatic hyperplasia, prior epidemiological studies only included men; thus, a pertinent question is does TZ similarly influence energy metabolism in men and women. Third, what is the dose-response relationship for TZ’s impact on energy metabolism in people? This point is relevant because TZ showed a biphasic effect on energy metabolism in mice, suggesting higher doses might counteract the potential energetic benefits.^34^ Lastly, what dose of TZ is both safe and effectively impacts energy metabolism in men and women? Women may be at a higher risk for developing orthostatic hypotension, highlighting the need to understand the impact of TZ on blood pressure changes in men and women.^41^ To address the above gaps, resolve the uncertainties surrounding the optimal dose of TZ, and pave the way for comprehensive studies and potential therapeutic breakthroughs in PD, we monitored the safety, tolerability, and bioenergetic target engagement of different doses of TZ in a mixed-sex cohort. Accordingly, we tested target engagement of TZ by evaluating the following parameters across escalating TZ doses: whole blood ATP levels corrected for hemoglobin (ATP/Hgb), ^31^Phosphorous-magnetic resonance spectroscopy (^31^P-MRS) to assay the ratio of βATP to inorganic phosphate (Pi) in the brain, ^18^F-fludeoxyglucose positron tomography (FDG-PET) to examine metabolic activity patterns, and plasma metabolomics.

## METHODS

### Participants

This study enrolled two cohorts of neurologically healthy older adults, aged between 60 and 85. Figure 1 displays the study timeline and titration schedules. Cohort A was comprised of six participants who were titrated to a daily TZ dose of 5 mg over five weeks. Cohort B included twelve participants who were titrated to 10 mg of TZ per day following a similar five-week schedule. The dosing regimens were designed based on the historical use of TZ and pharmacological considerations, with the higher dose in Cohort B providing insights into the effects of TZ at doses beyond the epidemiologically observed average.

**Figure 1.**
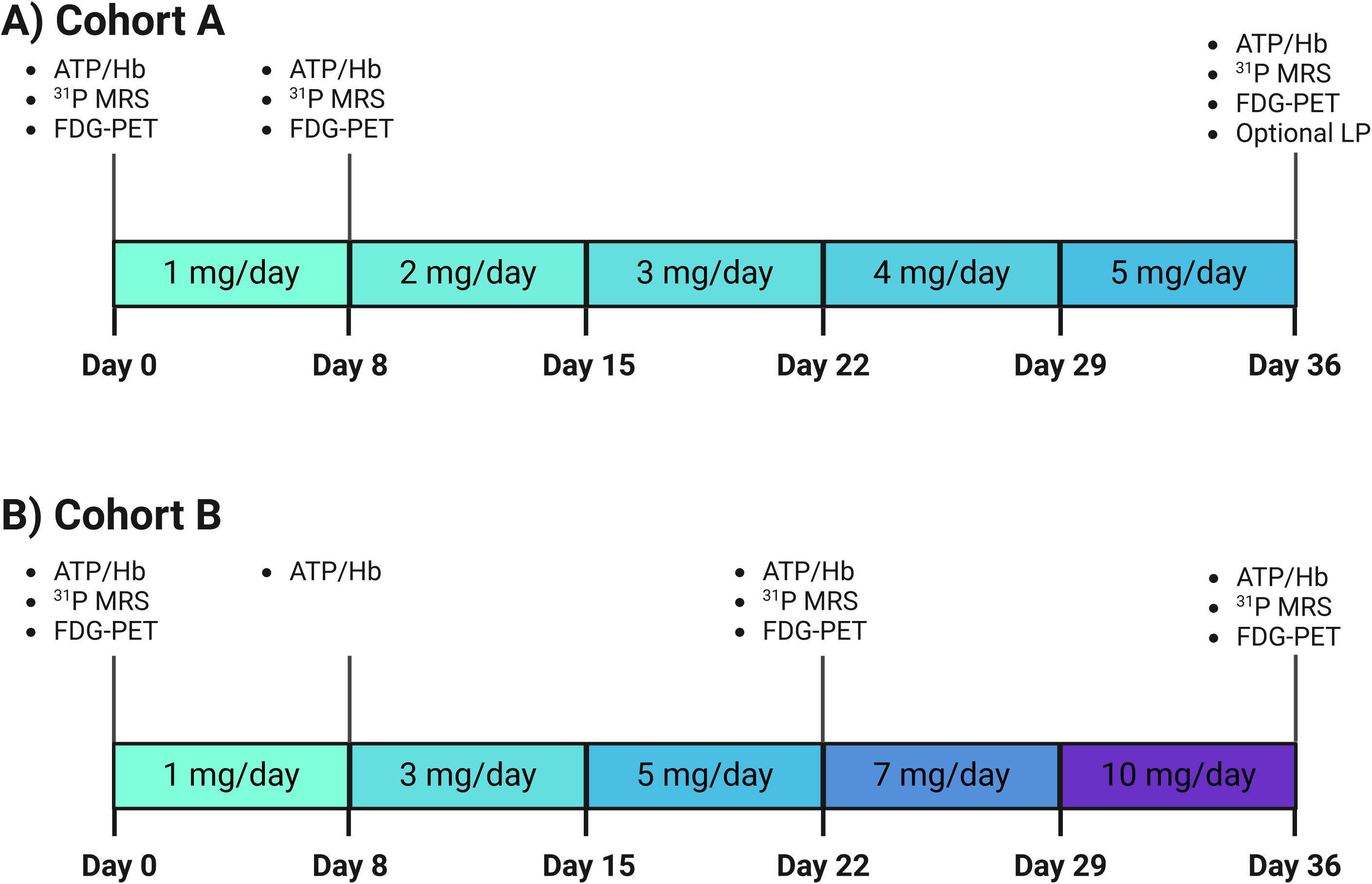
Study Design. **A)** There were six subjects in Cohort A. These subjects were titrated to a final dose of TZ of 5 mg/day over 5 weeks. Three subjects underwent a lumbar puncture after taking TZ 5 mg/day for one week. **B)** There were 12 subjects in Cohort B. these subjects were titrated to a final dose of TZ of 10 mg/day over 5 weeks. Subjects underwent four in-person visits which imaging studies occurring at baseline, when subjects were taking TZ 5 mg/day, and when subjects were taking TZ 10 mg/day.

Cohort A was sex-balanced (3 females and 3 males), had a mean age of 72.5 years, and an average Body Mass Index (BMI) of 24.3 kg/m^2^. Baseline blood pressure readings varied, with an average systolic pressure of 132 mmHg and a diastolic pressure of 83 mmHg. Orthostatic blood pressure and heart rate changes were also measured while supine and again after standing for three minutes. The mean change in systolic blood pressure (SBP) and diastolic blood pressure (DBP) was 1.8 mmHg and 6.2 mmHg, respectively, while the mean increase in heart rate (HR) was 10.7 beats per minute (bpm). Cohort B had a female majority (8 females and 4 males) with a mean age of 69.5 years, an average BMI of 27.3, and mean baseline blood pressure readings of 123/74 mmHg. The mean orthostatic changes were -4 mmHg for SBP, 4 mmHg for DBP, and 10 bpm for HR (Table 1). All participants were determined not to have PD or any other brain disease.

**Table 1:** Baseline Demographics. BMI: Body Mass Index HR: Heart rate N: Number rHR: Resting Heart Rate sDBP: Sitting Diastolic Blood Pressure sSBP: Sitting Systolic Blood Pressure S.D.: Standard Deviation

|  | Cohort A | Cohort B | p-value | All Subjects |
| --- | --- | --- | --- | --- |
| N | 6 | 12 | N/A | 18 |
| Age, mean ± S.D. | 73.0 ± 6.9 | 69.3 ± 8.6 | 0.38 | 70.5 ± 8.1 |
| % Male, n (%) | 3 (50.0) | 4 (33.3) | 0.86 | 7 (38.9) |
| BMI, mean ± S.D. | 25.9 ± 5.9 | 26.9 ± 3.6 | 0.65 | 26.6 ± 4.3 |
| rHR, mean ± S.D. | 65 ± 9.5 | 75 ± 11.9 | 0.09 | 72 ± 12.0 |
| sSBP, mean ± S.D. | 132 ± 11.5 | 123 ± 14.2 | 0.20 | 126 ± 13.8 |
| sDBP, mean ± S.D. | 83 ± 10.9 | 74 ± 7.3 | 0.07 | 77 ± 9.3 |
| Orthostatic $\Delta$ SBP, mean ± S.D. | 2 ± 5.6 | -4 ± 12.2 | 0.29 | -2 ± 10.6 |
| Orthostatic $\Delta$ DBP, mean ± S.D. | 6 ± 7.3 | 4 ± 8.3 | 0.54 | 5 ± 7.9 |
| Orthostatic $\Delta$ HR, mean ± S.D. | 11 ± 5.2 | 10 ± 7.1 | 0.80 | 10 ± 6.4 |

### Study Protocol

Participants in Cohort A began with a baseline assessment, including ^31^P-MRS, ^18^F-FDG PET, and a blood draw, before beginning a weekly titration of 1 mg TZ each week, starting at 1 mg (Fig. 1) and culminating in a final evaluation at 5 mg. The imaging and blood draws were performed after one week at 1 mg and again at 5 mg; the latter timepoint included an optional lumbar puncture, performed by University of Iowa neurologists using a sterile technique, undertaken by three volunteers. Cohort B followed a similar 5-week schedule that started with the same baseline assessments but a more accelerated titration (i.e., weekly 2 mg increases), starting from 1 mg and ending at 10 mg. Comprehensive follow-ups (i.e., ^31^P-MRS, ^18^F-FDG PET, and a blood draw) were performed after one week at 5 mg and again at 10 mg (Fig. 1).

Throughout the study, safety and tolerability were monitored, including orthostatic hypotension and potential drug-related adverse effects due to TZ’s known impact on blood pressure. All procedures and protocols were approved by the Institutional Review Board of the University of Iowa, and written informed consent was obtained from each participant before study initiation.

### Liquid chromatography-mass spectrometry (LC-MS) to measure TZ plasma and CSF concentration

The Metabolomics Core at the University of Iowa developed an assay to quantify TZ in the plasma and CSF using LC-MS.

#### Sample preparation and extraction

100 µl of plasma samples were mixed with 1290 µl of the chilled extraction solvent consisting of methanol, acetonitrile, and water in a 2:2:1 ratio (volume/volume/volume) containing 3.9 ng/ml of deuterium-labeled TZ. 10 µl of 100% water was added to the mixtures. Processed blank was prepared by adding only the extraction solvent to a microcentrifuge tube. Mixtures were vortexed for one minute and rotated at -20°C for 1 hour to precipitate protein. Next, samples were centrifuged at 21,000 x g for 10 minutes at 4LJC to remove insoluble material. 1000 µl of supernatants were transferred to clean microcentrifuge tubes and were evaporated to dryness using a Speed Vac and reconstituted in 20 µl of acetonitrile/water 1:1 ratio (v/v). Samples were stored at -20LJC for 2 hours and then centrifuged at 21,000 x g for 10 minutes at 4LJC. The supernatant was transferred to LC autosampler vials for analysis.

#### Calibration Curve

A standard dilution series was prepared in a commercially available human plasma sample (Innovative Research Inc, IPLAWBLIH50ML; Novi, MI) for TZ as described under sample preparation and extraction where 10 µl of TZ dilution was added in place of water. The final concentration of terazosin in the standard dilution series were 500, 250, 125, 62.5, 31.2,15.6 pg/µl. Samples for the calibration curve were prepared in triplicate. Three sample prepared without any spiked terazosin.

### Mass Spectrometry to quantify plasma metabolites

The Metabolomics Core at the University of Iowa used LC-MS and Gas Chromatography Mass Spectrometry (GC-MS) to quantify plasma metabolites.

#### LC-MS Analysis

LC-MS data were acquired on a Vanquish Flex UHPLC system (Thermo Scientific) coupled to a Thermo Q Exactive Hybrid Quadrupole-Orbitrap mass spectrometer. Two µL of metabolite extracts were separated using a Millipore SeQuant ZIC-pHILIC (2.1 × 150 mm, 5 µm particle size) column with a ZIC-pHILIC guard column (20 x 2.1 mm) attached to a Thermo Vanquish Flex UHPLC. Mobile phase was comprised of Solvent A: 20 mM ammonium carbonate, 0.1% ammonium hydroxide and Solvent B: acetonitrile. The mass spectrometer was operated in tSIM, positive polarity mode from 0 to 10 minutes, with the spray voltage set to 3.0 kV, the heated capillary held at 275 °C, and the HESI probe held at 350 °C. The sheath gas flow was set to 40 units, the auxiliary gas flow was set to 15 units, and the sweep gas flow was set to 1 unit. MS data acquisition was performed in a range of m/z 70–1,000, with the resolution set at 70,000, the AGC target at 5 × 104, and the maximum injection time at 200 ms. The chromatographic gradient during tSIM acquisition time was at a flow rate of 0.15 mL/min during tSIM data acquisition at linear gradient from 80 to 20% Solvent B.^42^

##### Data Processing

Xcalibur software (Thermo Fisher) was used for data acquisition and initial data quality control. TraceFinder software was used for peak analysis. Linear regression analysis of the standard dilutions series and extrapolation to unknowns was carried out in GraphPad Prism.

#### GC-MS

80 μL aliquots of plasma were also processed using GC-MS.^43^ Specifically, we were focused on quantifying pyruvate, the final metabolite in glycolysis, and citrate, a citric acid cycle metabolite that powers mitochondria. 80 μL of thawed plasma were extracted in an 18x volume to achieve a final acetonitrile:methanol:water ratio of 2:2:1, derivatized with Trimethylsilyl and methoxyamine, analyzed by a hard standard-verified (tier 1 confidence level) single quadrupole GC-MS (Thermo ISQ) method for relative quantitation of metabolites.

### Whole blood luminescence assay of ATP

Blood was collected in heparin anticoagulant and used for measurements of whole blood. Five microliters (µl) of whole blood were lysed in 45 µl detergent from the Abcam ATP detection assay kit (ab11384; Cambridge, UK) in triplicate. Samples were stored at -80°C until all blood draws were complete. Whole blood triplicates were thawed on ice and 10 µl of lysis was diluted into 30 µl of PBS. ATP measurements were performed with the CellTiter-Glo assay (Promega; Madison, WI). Each well contained 45 µl PBS, 50 µl CellTiter-Glo luciferase reaction and 5 µl of cell lysis. Samples were run on Spectramax L luminometer (Molecular Devices; San Jose, CA). The plates were covered, and the reaction was allowed to dark adapt for 10 minutes before reading each plate at 527 nm. Signals were compared to an ATP standard curve prepared in the same way. Hemoglobin was measured using a hemoglobin calorimetric assay kit (Cayman Chemical, cat # 700540; Ann Arbor, MI). Twenty µl of PBS diluted lysates was added to each well in a clear 96-well plate. One hundred eighty µl of 1x hemoglobin detection buffer was added to each well for a reaction volume of 200 µl. The plate was covered, mixed for 15 seconds and allowed to equilibrate for 15 minutes before absorbance measurements were taken at 575 nm. Sample values were compared to hemoglobin standards provided with the kit. Whole blood ATP levels were normalized to hemoglobin levels.^44^

### ^31^P Magnetic Resonance Spectroscopy

We acquired ^31^P-MRS spectra using an established protocol^40^ to quantify brain ATP fluctuations and evaluate target engagement. Participants undergoing the ^31^P MRS neuroimaging procedure were allowed to take their regular medications. Imaging was conducted using a GE SIGNA 7.0T MRI scanner equipped with a RAPID Biomedical ^31^P/^1^H dual-tuned birdcage head coil. Subjects were positioned within the coil for a series of scans, including 1) a localizer scan, 2) a ^1^H anatomical scan, 3) high-order B_0_ shimming, 4) Bloch-Siegert-based center frequency and transmitter gain optimization (PCr peak centered at 0 Hz), and 5) quantitative, non-localized ^31^P MRS scans. Automated high-order shimming of the whole brain was performed with deep-learning-based autoHOS software.^45^ The ^31^P MRS data collection employed a free induction decay (FID) sequence with a short block-shaped excitation pulse (152 μs) at a 20° flip angle. Each scan consisted of 128 acquisitions, with parameters including a TR of 2000 ms, 2048 sampling points, and a spectral width of 10,000 Hz. Three FID replicates at each time point were acquired to assess measurement uncertainty. FIDs were zero-filled to 8K and apodized with 5 Hz line-broadening prior to Fourier-Transformation. Spectral analyses were performed on magnitude spectra for enhanced reproducibility. Metabolite peaks were identified within fixed boundaries (PCr: -1.5 to -2.0 ppm, αATP: -9.0 to -6.0 ppm, βATP: -18.0 to -14.0 ppm, γATP: -4.0 to -1.5 ppm, and inorganic phosphate: 4.4 to 5.8 ppm), with peak areas integrated as the summation of data points within these ranges. This approach accounted for minor peak position shifts due to pH-induced variations. Peak positions were determined by locating the maximum spectral amplitude. Total spectral signal intensity was computed by summing all data points within the -25 ppm to +25 ppm range.

### ^18^F-FDG PET Imaging

Participants were instructed to undertake a calorie fast for at least 6 hours before FDG injection. Blood glucose was checked via finger stick and glucose levels were required to be ≤ 200 mg/dl to proceed with imaging. Scanning was performed a GE Discovery MI Time of Flight PET/CT camera with SiPM array detector technology (GE Healthcare, Waukesha, WI). Participants were laid supine on the imaging table and imaging commenced when 5 mCi of FDG was administered via venous catheter. The participant rested on the imaging table with eyes open and ears unplugged for the duration of the 60 min dynamic image collection. Imaging was dynamically collected in six frames of 5 s each (i.e., 6 x 5 s) followed by 9 x 10 s, 10 x 30 s, 13 x 60 s, and 8 x 300 s beds (3600 s = 60 min total). Computed Tomography (CT) images were acquired immediately before the PET scan and were utilized for attenuation-correction of the ensuing PET image. All imaging sets were reconstructed using VUE Point HD reconstruction with 16 subsets and 6 iterations; PET images were also corrected for tracer decay, attenuation, scatter, and dead-time. For Cohort A, ^18^F-FDG PET was acquired at baseline, after one week at 1 mg TZ, and after one week at 5 mg TZ. Cohort B was imaged at baseline, after one week at 5 mg TZ, and after one week at 10 mg TZ. Structural T1-weighted MPRAGE brain magnetic resonance imaging (MRI; used for PET co-registration during analysis) was acquired on a GE Discovery 950 7T scanner with a 32-channel head coil at 1 mm isotropic voxel size (Cohort A) and 0.43 x 0.43 x 0.69 mm voxel size (Cohort B). For co-registration with PET, all Cohort B T1-weighted images were resampled to 1mm isotropic in PMOD (PMOD Technologies LLC, Zurich, Switzerland) using SINC interpolation.

PET image preprocessing and analysis were performed with PMOD version 4.4 modules. The final 30 min of the 60-min scan were spatially aligned (i.e., motion corrected) and then summed to create a static ^18^F-FDG PET image. Using the Brain Parcellation workflow in the PNEURO module, the structural MRI was segmented into gray and white matter and probabilistically parcellated into cortical and deep nuclei volumes of interest (VOIs) according to Hammers N30R83 atlas. The static PET image was then co-registered with the structural MRI and the average standardized uptake value (SUV) of each VOI was calculated. A volume-weighted global mean comprised of all VOIs, excluding cerebrospinal fluid and white matter regions, was also calculated to represent systemic (i.e., whole-brain) metabolic changes.

### Statistical Analysis

For all analyses, we constructed linear mixed-effect regression analyses with a random effect per subject to evaluate changes in outcomes as a function of dose. For analyses of changes in blood pressure from baseline, we controlled for baseline age, sex, and BMI. These same covariates were included in the analysis of the concentration of TZ at varying doses. The models that evaluated the percent change from baseline in whole blood ATP and change in FDG uptake included the covariates of baseline age, sex, BMI, and baseline ATP or baseline FDG uptake, respectively. These models were repeated to evaluate the dose-by-sex interaction to determine if there were significantly different dose effects as a function of sex.

All analyses were performed in RStudio version 4.3.1 using the lmerTest package. Unless otherwise stated, individual data points in all figures represent unadjusted values, while indicators of aggregated data, such as means with error bars, represented the predicted mean from the models after adjusting for covariates. Results were considered significant if the p-value was < 0.05.

## RESULTS

### Protocol

The baseline demographic characteristics of the participants are shown in Table 1. We screened a total of 19 participants. One withdrew prior to the baseline visit, leaving six in Cohort A and 12 in Cohort B. One participant from Cohort B was unable to complete the final study visit due to severe weather. All other participants completed all aspects of the study and provided full datasets.

### TZ concentration in plasma and cerebral spinal fluid

As the TZ dose increased, the plasma TZ concentration increased approximately linearly. The TZ concentration at 1 mg/day was 0.02±0.01 ng/mL (mean ±SEM), at 5 mg/day was 0.08±0.01 ng/mL, and at 10 mg/day was 0.14±0.01 ng/mL (Supp. Fig. 1). The concentration of TZ was significantly higher relative to baseline at all doses (p<0.01). These changes did not differ significantly as a function of sex (Supp. Table 1).

To test TZ brain penetration, we obtained cerebral spinal fluid (CSF) by lumbar puncture in three volunteers from Cohort A who took TZ 5 mg/day for 1 week. We measured plasma and CSF TZ concentrations and calculated the plasma-free TZ concentration (TZ is ∼92% protein-bound^46^; Supp. Figure 2). The CSF TZ concentration was 23±3 % (mean ± SEM) of the free plasma concentration, a ratio expected based on plasma availability (Supp. Figure 2).^46^

### ATP/Hb levels in whole blood

A TZ dose of 5 mg/day increased ATP/Hb levels in whole blood by 6.2±1.7% (mean ±SEM) compared to baseline values (p<0.01) and to 1 mg/day (p=0.006; Fig 2). Moreover, ATP/Hb levels at the 5 mg/day dose increased >5% in 11 of the 18 subjects. The increases from baseline at TZ 1 mg/day and 10 mg/day were not statistically significant compared to baseline (p=0.46 and p=0.06, respectively; Fig. 2). There were no differences as a function of sex (Supp. Table 1).

**Figure 2.**
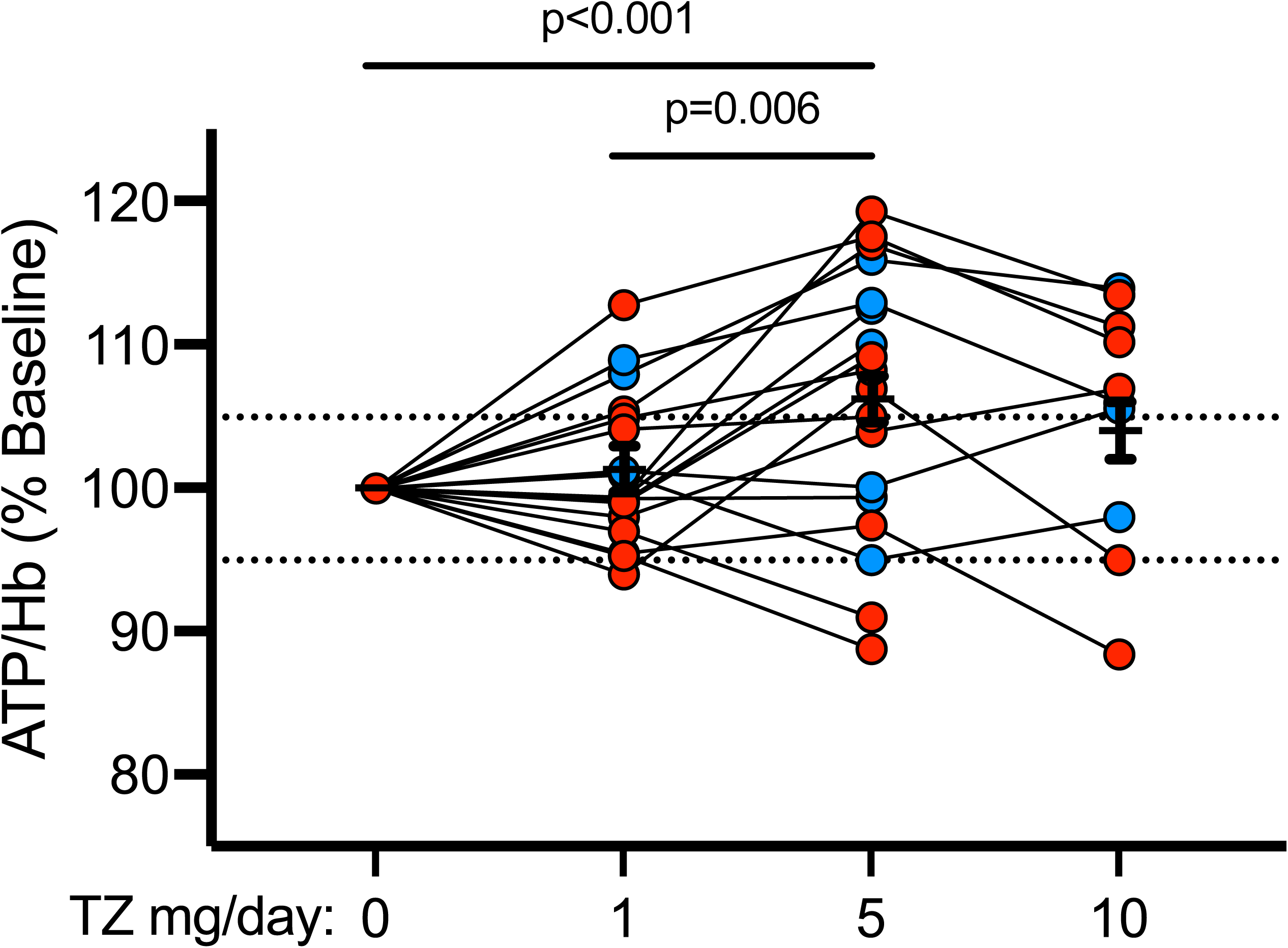
Changes in ATP/Hb by TZ Dose. There was not a statistically significant increase from baseline in ATP/Hb at 1 mg/day of TZ (1.26% ± 1.62; mean ± SEM, p=0.46). However, ATP/Hb was significantly elevated from baseline at 5 mg/day (6.22% ± 1.62; p<0.01). ATP/Hb was increased by 4.10% ± 2.02 (mean ± SEM) at 10 mg/day, but this was not statistically significant (p=0.06). TZ 5 mg/day was associated with a significantly higher increase in ATP/Hb compared to 1 mg/day (p<0.01). TZ 10 mg/day was not significantly different from TZ 5 mg/day (p=0.32). Dashed lines represent a 5% increase or decrease and presented for visualization purposes only. Data were available from 18 subjects at TZ doses of 1 mg/day and 5 mg/day. Data were available for 10 subjects at TZ 10 mg/day.

### Ratio of ***β***ATP/P_i_ in brain

We used ^31^P-magnetic resonance spectroscopy to assay the ratio of βATP to inorganic phosphate (βATP/P_i_) in brain. The changes from baseline (mean ±SEM) at a TZ dose of 1 mg/day were 0.50±1.90% (n=6, p=0.80), at 5 mg/day were 0.19±1.35 % (n=16, p=0.89), and at 10 mg/day were -1.64±1.63% (n=9, p=0.32). There were no differences as a function of sex (Supp. Table 1).

### ^18^F-FDG PET in brain

We found that TZ 5 mg/day decreased the global FDG uptake in the brain (-6.14±2.15%) (mean ± SEM) (p<0.01; Fig. 3). TZ 10 mg/day was also associated with a slightly greater decrease in FDG uptake (-6.59±2.58%; p=0.01), but this was not more significant than 5 mg/day (p=0.86). TZ 1 mg/day did not significantly change FDG uptake from baseline (-1.36±3.06%; p=0.66). There was no difference in FDG uptake as a function of sex (Supp. Table 1).

**Figure 3.**
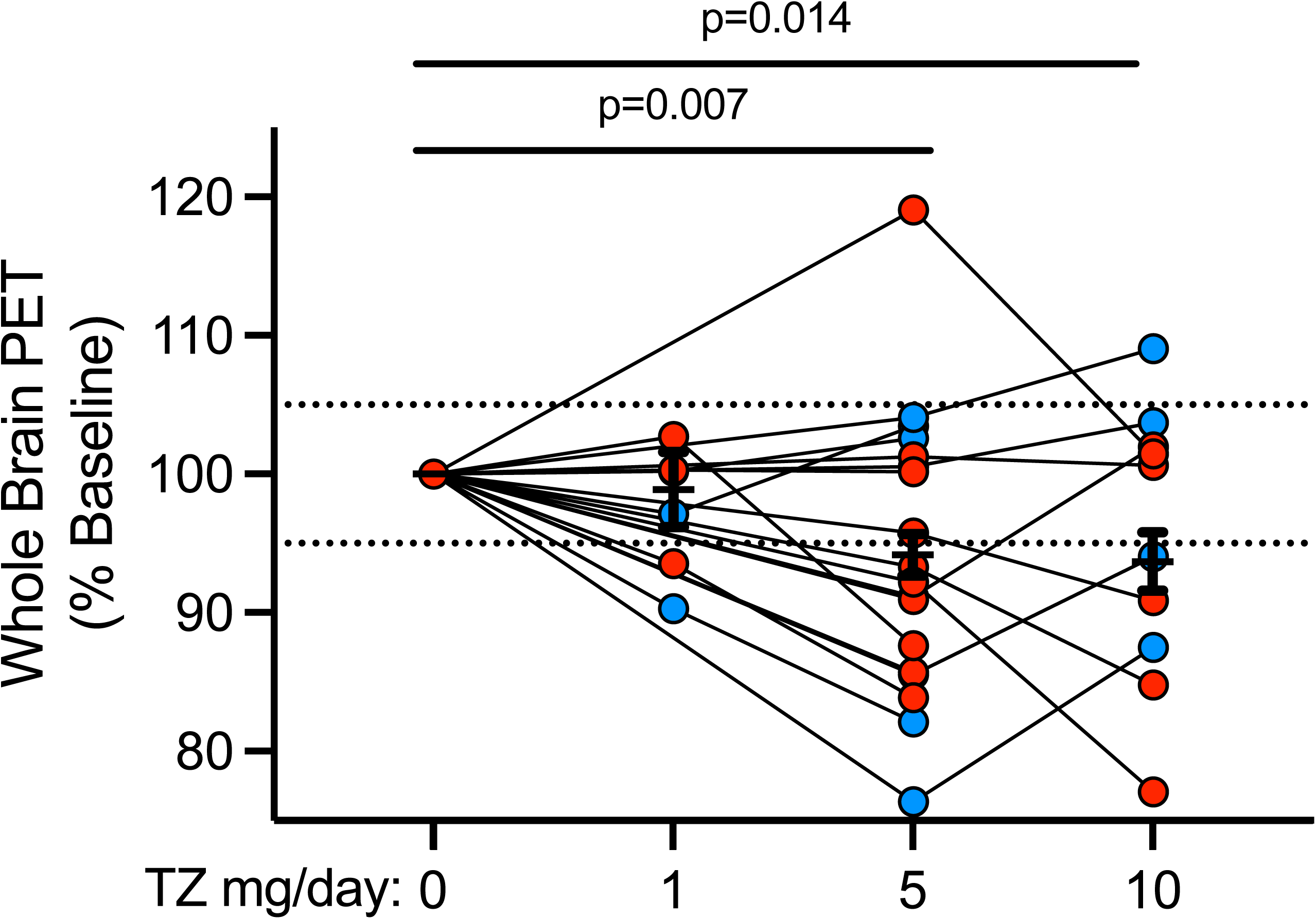
Changes in Brain 18F-FDG PET Signal by TZ Dose. The global standard uptake value from the 18F-FDG PET studies showed a significant decrease in signal from baseline at TZ doses of 5 mg/day (mean = -6.14%, SEM = 2.15, p<0.01) and 10 mg/day (mean = -6.59, SEM = 2.58, p=0.01). TZ 5 mg/day was not significantly different from TZ 1 mg/day (p=0.13) or TZ 10 mg/day (p=0.86). The mean decrease at a dose of TZ 1 mg/day was -1.36 ± 3.06 (mean ± SEM; p=0.66).

Importantly, the decrease in ^18^F-FDG PET signal was a universal phenomenon. Specifically, the global ^18^F-FDG PET signal was the average of the SUV from various regions. Therefore, key regions, such as the basal ganglia, may have shown an increase in ^18^F-FDG PET uptake.

However, we saw a universal decrease in all regions-of-interest, including the basal ganglia and substantia nigra.

### Plasma GC-MS Metabolomic Analysis

When evaluating changes in the concentration of pyruvate, one data point was removed for being more than three standard deviations from the mean. We found that TZ 1 mg/day increased plasma pyruvate concentrations from baseline (61.28±17.44%; p<0.01). The difference in pyruvate concentration from baseline at TZ 5 mg/day was 17.51±17.74 (p=0.33) and at TZ 10 mg/day was 12.26±21.31 (p=0.57). There were no differences in concentrations of pyruvate as a function of sex at any doses (Supp. Table 1).

We found that TZ 5 mg/day increased plasma citrate concentrations from baseline (19.06±6.31%; p=0.03) (mean ± SEM) (Supp. Fig. 3). TZ increased plasma citrate at doses of 1 mg/day (15.26±6.31%) and 10 mg/day (16.28±8.62%), but the results did not reach statistical significance (p=0.09 and p=0.14, respectively). There were no differences in citrate plasma concentration between TZ 1 mg/day and TZ 5 mg/day, nor was there a difference between TZ 5 mg/day and TZ 10 mg/day. There were no differences as a function of sex in citrate concentrations (Supp. Table 1).

### Safety and Tolerability

An older female participant in her 80’s from Cohort B withdrew when she reached a dose of 7 mg TZ because of dizziness and lightheadedness. Two female participants in Cohort B reported mild dizziness at a dose of 5 mg/day TZ. One male participant in Cohort B reported mild dizziness at a dose of 10 mg/day TZ. We could discern no relationship between the report of dizziness and a history or treatment of hypertension. These events were classified as mild and did not necessitate discontinuation of the study. There were no other adverse events.

### Blood pressure

Table 1 shows baseline systolic (SBP) and diastolic blood pressures (DBP). SBP and DBP decreased significantly from baseline at all doses of TZ (Supp. Table 2), consistent with its α-1 adrenergic receptor antagonistic activity. The mean decrease in SBP and DBP were similar at TZ doses of 5 mg/day and 10 mg/day (Supp. Fig. 4A-B). There were no significant sex differences in the observed changes in vital signs at each dose (Supp. Table 1).

We also measured orthostatic changes in blood pressure and heart rate at all study visits. There were no significant orthostatic changes in SBP or DBP at any dose of TZ; however, there was a significant increase in orthostatic heart rate at TZ 5 mg/day (Supp. Table 2; Fig. 5A-C). There were no significant differences in the orthostatic values as a function of sex (Supp. Table 1).

### Post-Hoc Analysis

After observing a significant decrease in FDG uptake in the TZ 5 mg/day group, we hypothesized that this may represent an increase in glycolytic efficiency. Specifically, glucose is the primary input of glycolysis, and if TZ is promoting glycolytic efficiency this would manifest as decreased glucose uptake (i.e., decreased FDG uptake). To test this post-hoc hypothesis, we evaluated the relationship between the glycolytic by-product of pyruvate and FDG uptake. This was performed using all data points from each subject using a linear mixed effects regression analysis that included a per-subject random effect. There was a significant, negative relationship between the percent change from baseline of FDG uptake and the percent change from baseline in pyruvate (p=0.04). Specifically, as FDG uptake decreased, plasma pyruvate concentrations increased (Supp. Fig. 6).

## DISCUSSION

The purpose of this study was to assess the safety, tolerability and bioenergetic target engagement of different TZ doses in a mixed-gender cohort. Our findings suggest that TZ 5 mg/day might represent the best balance between efficacy and safety/tolerability. Indeed, the 5 mg/day dose effectively reached its primary target to improve energy metabolism in healthy controls, was well tolerated, and equally impacted both men and women. These data suggest a potential neuroprotective benefit at a TZ dose of 5 mg/day which bears rigorous study in clinical trials.

This study provides insight into the relationship between the dose/concentration of TZ and its metabolic effects. For both ATP/Hb levels in whole blood and brain FDG uptake, a dose of 1 mg/day TZ did not significantly alter values, while 5 mg/day yielded significant differences and 10 mg/day did not generate further changes. These results are consistent with findings that the relationship between TZ concentration/dose and the activity of PGK1 as an isolated protein, in cells and in animals, is biphasic; low TZ concentrations increase activity and high concentrations inhibit PGK1.^34^ The biphasic response is an inherent property of the TZ:PGK1 interaction and we recently provided a molecular explanation for this unusual behavior.^47^

Neuroimaging with FDG-PET has previously revealed metabolic dysfunction in PD patients.^48^ Our data indicate that TZ decreased brain FDG uptake and our post-hoc analysis demonstrated a significant, negative correlation between pyruvate and FDG-PET in support of the notion that the TZ-induced decrease in FDG uptake was the result of increased glycolytic efficiency. Finding decreased FDG uptake throughout the brain might be expected for a drug that non-selectively affects PGK1 in all cells and in a study of healthy controls without neurodegenerative disease. These results suggest that FDG-PET may be a valuable assay of TZ target engagement in future studies.

In a previous pilot study, we found that 5 mg/day of TZ increased βATP/Pi in the brain as measured by ^31^P-MRS.^40^ However, this result was not replicated in this study. One potential reason is that TZ might more strongly affect βATP/Pi levels in PD patients who have decreased ATP^49–51^, but not in neurologically healthy controls who have presumably normal ATP levels. The mechanism of TZ action and these considerations suggest that clinical trials of TZ would also benefit by including assays of βATP/Pi to assess target engagement.

In addition to its desired target in PD (PGK1), TZ inhibits the α-1 adrenergic receptor, which raises the possibility that a decrease in blood pressure could generate adverse effects. In this study, one participant withdrew because of dizziness and three others reported mild dizziness not leading to withdrawal. Notably three of those four were females, and the male who reported dizziness was taking the highest, 10 mg/day dose. In another study of people with PD, three (all females) of the eight participants stopped taking TZ because of dizziness and/or orthostatic hypotension.^40^ Taken together, these results raise the question of whether there is a differential effect of α-1 adrenergic receptor inhibition in women. Interestingly, in a large epidemiological study, males with PD who were taking TZ had a decrease, rather than an increase, in diagnoses that might be attributed to hypotension.^34^ In any case, these data, plus the presence of autonomic insufficiency in some people with PD^52^, warrant careful attention to this potential adverse event in trials of TZ in PD.

A limitation of this study is that we used neurologically healthy participants and dose-response relationships, and adverse events may differ in PD patients who have unique biological features. On the other hand, our data may have applicability for additional neurodegenerative diseases that also exhibit impaired brain bioenergetics, like those that have reported TZ efficacy in models of Alzheimer’s disease^53^, amyotrophic lateral sclerosis^54^, and spinal muscular atrophy.^55^ Another limitation is the short duration of this study. Still, these findings suggest that TZ has a favorable safety profile in long-term use, including benefits in several large epidemiological studies.^35–39^

Our findings in neurologically healthy participants suggest a potential benefit of 5 mg/day TZ for slowing or preventing neurodegeneration in energetically deficient populations, like people with PD. When combined with other results in PD patients^40^, they also suggest that clinical trials aiming to test this supposition will benefit from including multiple methods of target engagement, including blood ATP/Hb, brain ^18^F-FDG PET, and βATP/Pi via ^31^P-MRS. Measuring target engagement in a PD clinical trial would provide the unique opportunity to correlate clinical and biochemical changes resulting from TZ and provide insight into fundamental mechanisms.

## Data Availability

All data produced in the present study are available upon reasonable request to the authors.

## ACKNOWLEDGEMENTS

This study was funded by the Michael J. Fox Foundation for Parkinson’s Research (to JLS and NSN), NIH Grant 5UL1TR002537-04 through the University of Iowa Institute for Clinical and Translational Science and was conducted using equipment supported by NIH Grant S10RR028821, and NIH Grant K23NS117736 (to JLS). MJW is an investigator of the Howard Hughes Medical Institute. The study sponsors had no role in the conduct, analysis, or reporting of these results.

## AUTHOR CONTRIBUTIONS

(1) Research project: (a) conception, (b) organization, (c) execution. (2) Statistical analysis: (a) design, (b) execution, (c) review and critique. (3) Manuscript preparation: (a) writing of the first draft, (b) review and critique. JLS: 1abc, 2abc, 3ab; PEG: 1bc, 2abc, 3ab; CDW: 1bc, 2abc, 3ab; LLP: 1bc, 2abc, 3ab; SC: 1bc, 3ab; CSN: 1c, 3ab; CLG: 1c, 3b; EBT: 1ac, 2ab, 3b; SEE: 1abc, 3ab; JX: 1abc, 2bc, 3ab; EYU: 1a, 3ab; VAM: 1abc, 2b, 3ab; MJW: 1ab, 2abc, 3ab; NSN: 1abc, 2abc, 3ab.

## ADDITIONAL INFORMATION

Correspondence and requests for materials should be addressed to JLS or NSN.

