## Supplemental Material for "A pilot dose-finding study of Terazosin in humans"

**Supplemental Table 1: Change from Baseline as a Function of Sex**

| Outcome Measure | TZ Dose | Males relative to Females  Estimate ± SEM | p-value |
| --- | --- | --- | --- |
| SBP  (mmHg) | 1 mg/day | -7 ± 5.53 | 0.25 |
|  | 5 mg/day | -1 ± 5.53 | 0.82 |
|  | 10 mg/day | -13 ± 6.78 | 0.07 |
| DBP  (mmHg) | 1 mg/day | -2 ± 3.35 | 0.49 |
|  | 5 mg/day | 3 ± 3.35 | 0.39 |
|  | 10 mg/day | -3 ± 4.11 | 0.46 |
| Orthostatic SBP  (mmHg) | 1 mg/day | 4 ± 6.86 | 0.56 |
|  | 5 mg/day | 3 ± 6.86 | 0.65 |
|  | 10 mg/day | 1 ± 8.32 | 0.93 |
| Orthostatic DBP  (mmHg) | 1 mg/day | 5 ± 4.52 | 0.25 |
|  | 5 mg/day | 7 ± 4.52 | 0.14 |
|  | 10 mg/day | 7 ± 5.40 | 0.24 |
| Orthostatic HR  (beats per minute) | 1 mg/day | -4 ± 3.43 | 0.21 |
|  | 5 mg/day | 1 ± 3.43 | 0.72 |
|  | 10 mg/day | 1 ± 4.19 | 0.87 |
| Plasma [TZ]  (μg/mL) | 1 mg/day | -0.01 ± 0.01 | 0.88 |
|  | 5 mg/day | -0.01 ± 0.01 | 0.84 |
|  | 10 mg/day | -0.01 ± 0.016 | 0.51 |
| Whole Blood ATP  (LU/mg) | 1 mg/day | 1.94 ± 3.66 | 0.60 |
|  | 5 mg/day | 0.74 ± 3.66 | 0.84 |
|  | 10 mg/day | 2.46 ± 4.46 | 0.58 |
| βATP/Pi  (Arbitrary Units) | 1 mg/day | 4.79 ± 3.88 | 0.23 |
|  | 5 mg/day | 1.05 ± 2.78 | 0.71 |
|  | 10 mg/day | 0.87 ± 3.36 | 0.80 |
| Whole Brain  FDG-PET  (SUV) | 1 mg/day | -6.52 ± 6.03 | 0.29 |
|  | 5 mg/day | -1.24 ± 4.30 | 0.77 |
|  | 10 mg/day | 7.84 ± 5.10 | 0.13 |
| Citrate Plasma Concentration  (Arbitrary Units) | 1 mg/day | 13.37 ± 17.87 | 0.46 |
|  | 5 mg/day | 11.10 ± 17.87 | 0.54 |
|  | 10 mg/day | -0.63 ± 22.07 | 0.98 |
| Pyruvate Plasma Concentration  (Arbitrary Units) | 1 mg/day | 39.94 ± 36.60 | 0.28 |
|  | 5 mg/day | 43.29 ± 37.89 | 0.26 |
|  | 10 mg/day | 9.89 ± 45.22 | 0.83 |

**Supplemental Table 2: Blood Pressure and Heart Rate Changes from Baseline**

| Outcome Measure | TZ Dose | Change Relative to Baseline  Estimate ± SEM | p-value |
| --- | --- | --- | --- |
| SBP  (mmHg) | 1 mg/day | -6 ± 2.7 | 0.02 |
|  | 5 mg/day | -13 ± 2.7 | <0.01 |
|  | 10 mg/day | -11 ± 1.6 | <0.01 |
| DBP  (mmHg) | 1 mg/day | -3 ± 1.6 | 0.04 |
|  | 5 mg/day | -10 ± 2.0 | <0.01 |
|  | 10 mg/day | -9 ± 4.11 | <0.01 |
| Orthostatic SBP  (mmHg) | 1 mg/day | -1 ± 3.3 | 0.80 |
|  | 5 mg/day | 2 ± 2.3 | 0.54 |
|  | 10 mg/day | -6 ± 4.0 | 0.15 |
| Orthostatic DBP  (mmHg) | 1 mg/day | -2 ± 2.2 | 0.38 |
|  | 5 mg/day | 0 ± 2.2 | 0.97 |
|  | 10 mg/day | -5 ± 2.6 | 0.08 |
| Orthostatic HR  (beats per minute) | 1 mg/day | 1 ± 1.7 | 0.45 |
|  | 5 mg/day | 5 ± 1.7 | 0.02 |
|  | 10 mg/day | 1 ± 2.1 | 0.68 |

**Supplemental Figure 1: TZ Plasma Concentration by Dose**

The concentration of TZ was significantly higher from baseline at each subsequent dose increase. Additionally, TZ plasma concentration was significantly different from all doses at a level of p<0.01. Data were available for 18 subjects at TZ 1 mg/day, 18 subjects at TZ 5 mg/day, and 10 subjects at TZ 10 mg/day.

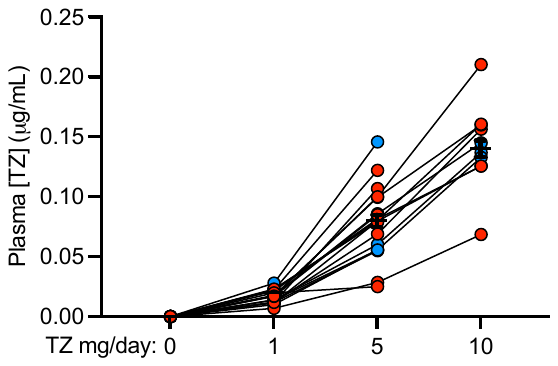

**Supplemental Figure 2: TZ Concentration in CSF**

TZ was reliably detected in the CSF of all the three subjects from Cohort A who underwent a lumbar puncture while taking a TZ dose of 5 mg/day.

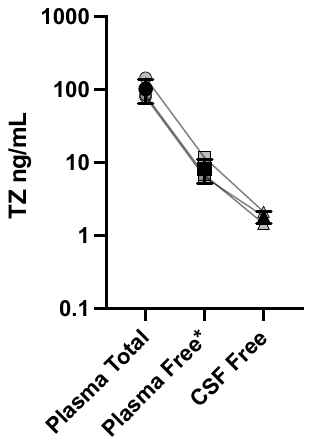

**Supplemental Figure 3: Changes in Plasma Citrate by TZ Dose**

The concentration of plasma citrate was not significantly elevated from baseline at TZ 1 mg/day (15.26±6.31%, p=0.89), but it was elevated at TZ 5 mg/day (19.06±6.31 %, p=0.03). At TZ 10 mg/day, plasma citrate increased by 16.28±8.62% (p=0.14).

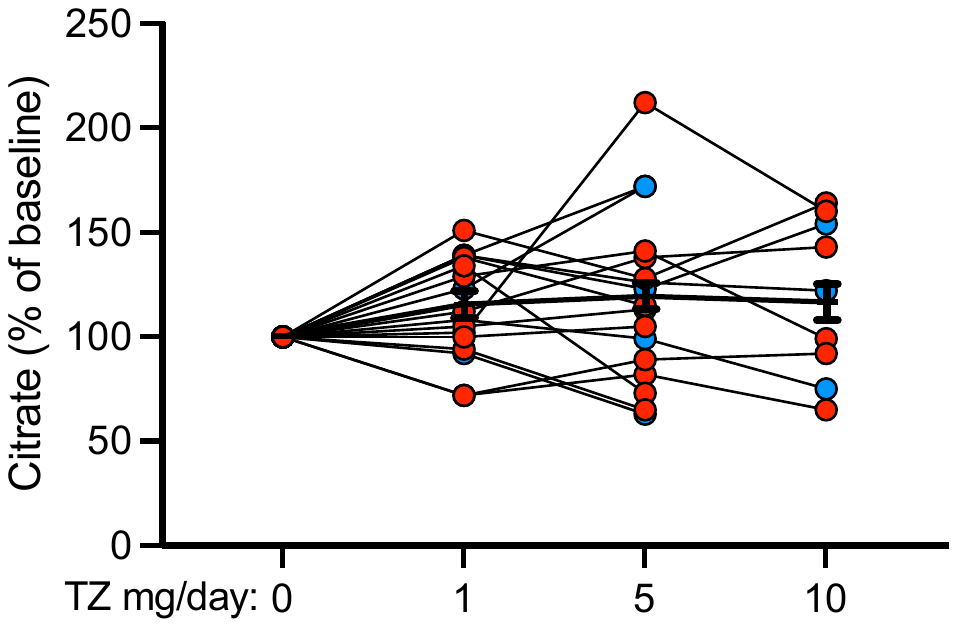

**Supplemental Figure 4. Changes in Blood Pressure by TZ Dose**

**a-b)** Systolic Blood Pressure decreased from baseline at all doses of TZ. Blood pressure lowering at 5 mg/day was twice that seen as 1 mg/day. There were no differences in blood pressure lowering between 5 mg/day and 10 mg/day. Further details are outlined in Supplemental Table 2.

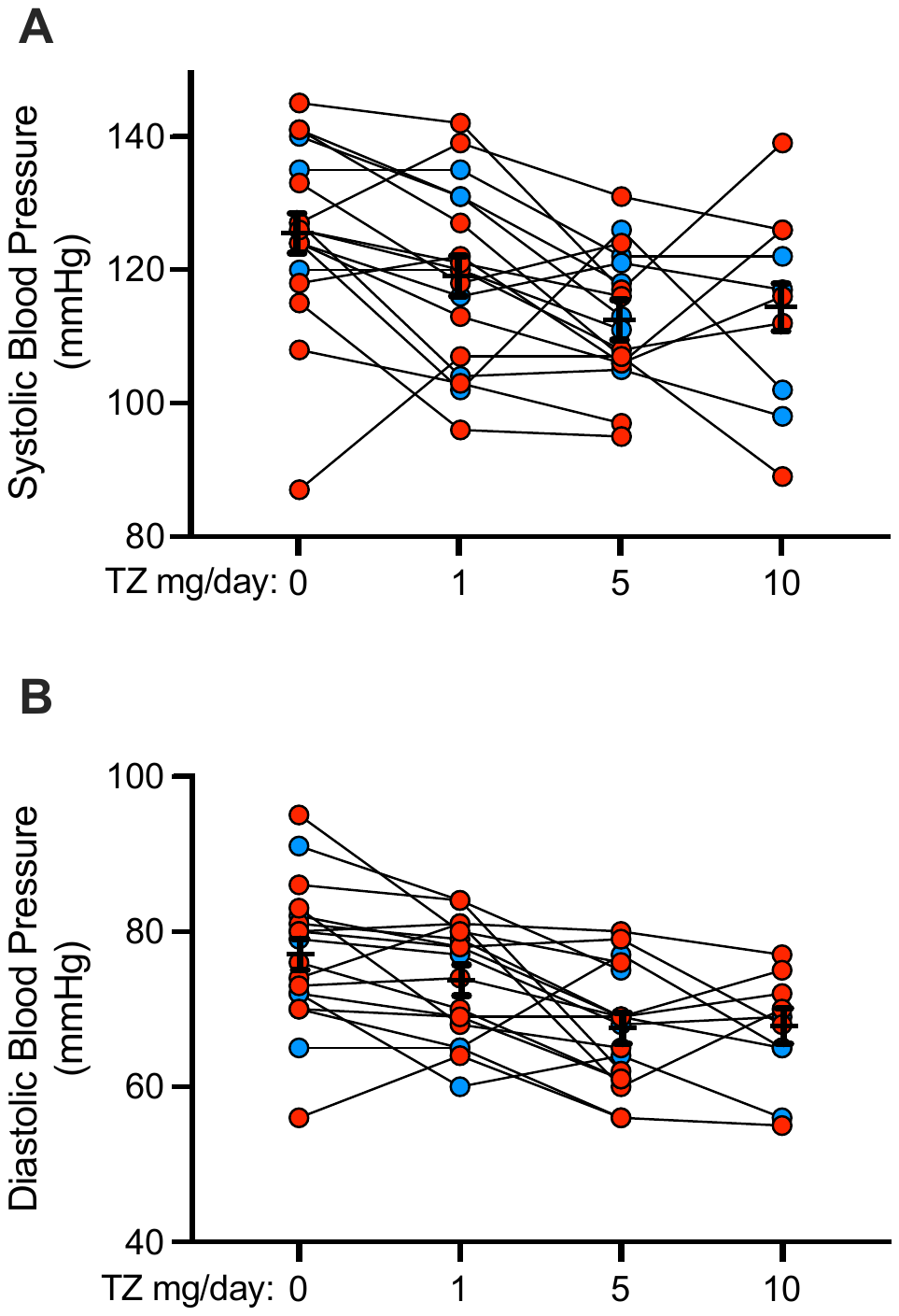

**Supplemental Figure 5: Orthostatic Changes by TZ Dose**

**a-c)** Orthostatic changes in blood pressure and heart rate did not differ significantly from baseline at any dose of TZ. Further details are outlined in Supplemental Table 2.

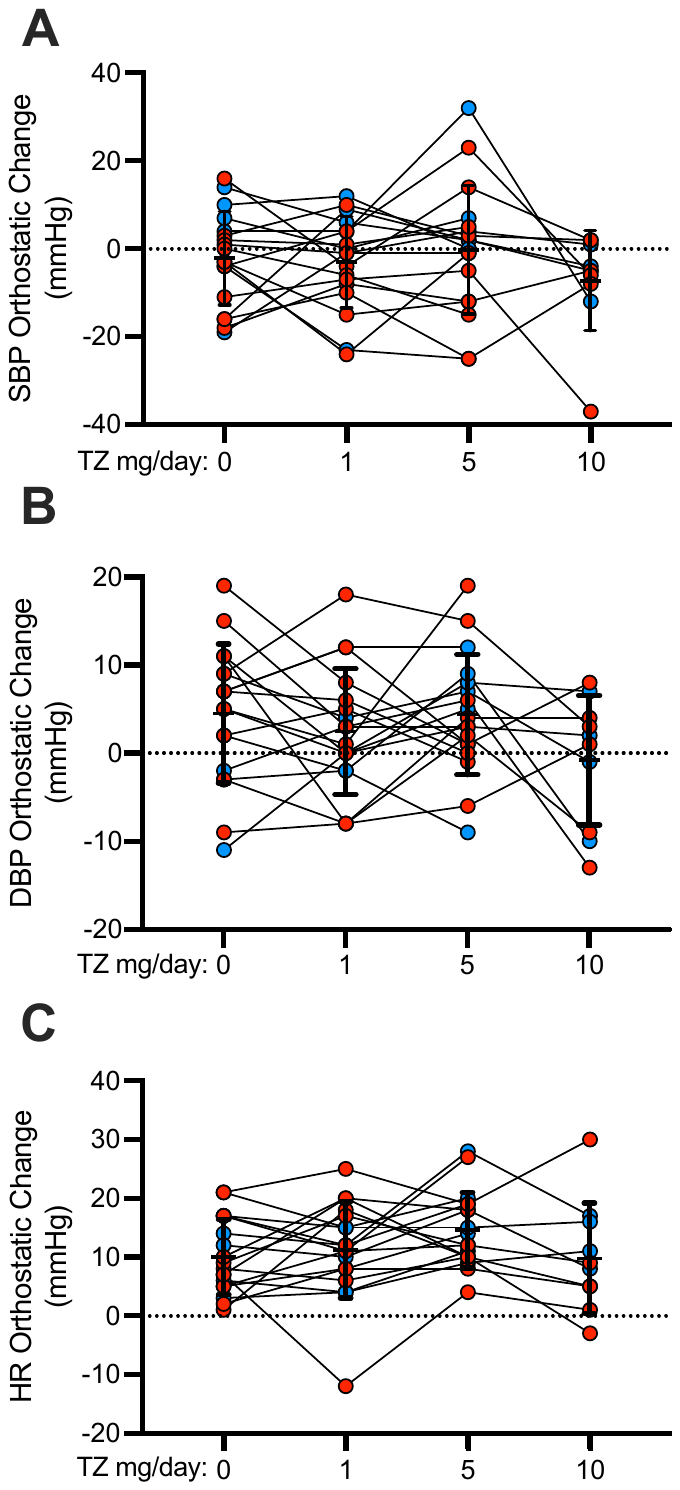

**Supp. Figure 6: Changes in Brain FDG-PET Signal Correlate with Changes in Plasma Pyruvate Concentration**

Our post-hoc analysis demonstrated a significant, negative relationship between the decrease in FDG-PET uptake signal from baseline and the change in plasma pyruvate concentration from baseline. These findings indicate that a decrease in FDG-PET signal may be indicative of improved glycolytic efficiency.

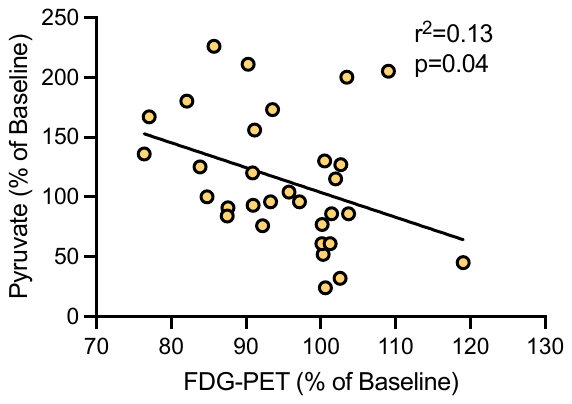
